# Biomechanical Analysis of Dynamic Gripping in Manual Laborers Exhibiting Work-Related Scapholunate Instability Signs

**DOI:** 10.64898/2026.06.27.26356743

**Authors:** Muhammad Hassan Bari, Zaira Khalid, Moazma Ilyas, Sharia Qaiser

**Author notes:** **Corresponding Author:** Muhammad Hassan Bari, Department of Physical Therapy, Islam College of Physical Therapy, Sialkot, Pakistan.

## Abstract

**Background:** Scapholunate instability (SLI) represents the most prevalent form of carpal instability and is increasingly recognized as a clinically significant occupational condition in manual-labor populations. Repetitive forceful gripping, sustained wrist loading, and awkward joint postures inherent to construction, manufacturing, and heavy industry may predispose workers to progressive scapholunate ligament compromise; however, the specific biomechanical mechanisms underlying dynamic grip force transmission in symptomatic laborers remain poorly characterized.

**Objective:** This study aimed to quantify dynamic grip force profiles and wrist kinematics in manual laborers exhibiting clinical signs of scapholunate instability; compare these biomechanical parameters with asymptomatic worker controls; and identify occupation-specific loading patterns that may contribute to scapholunate ligament compromise.

**Methods:** A cross-sectional observational design was employed. Forty-two male manual laborers, mean age 36.4 (SD 7.2) years; mean occupational exposure 9.8 (SD 4.1) years, were recruited from construction and surgical instrument manufacturing sites in Sialkot District, Pakistan. Participants were stratified into a symptomatic group (n = 21) based on positive Watson scaphoid shift test, dorsal wrist pain, and functional limitation, and an asymptomatic control group (n = 21). Dynamic grip force was measured using a calibrated Jamar dynamometer across five standardized trials. Wrist kinematics were captured during a simulated gripping task using electrogoniometry. Surface electromyography (sEMG) recorded flexor digitorum superficialis, flexor carpi ulnaris, and extensor carpi radialis longus activity. Scapholunate gap was confirmed radiographically. Statistical analyses included independent samples t-tests, Pearson correlation, and logistic regression (α = 0.05).

**Results:** Symptomatic workers demonstrated significantly reduced grip force (28.4 (SD 5.6) kg versus 41.7 (SD 6.3) kg; *p* < 0.001) and elevated wrist ulnar deviation during peak gripping (22.3 (SD 4.1) degrees versus 14.6 degrees (SD 3.8) degrees; *p* = 0.002) compared with controls. sEMG amplitude of flexor digitorum superficialis was paradoxically elevated in the symptomatic group (mean difference 18.3 microvolts; 95% CI 11.4-25.2), suggesting compensatory hyperactivation. Scapholunate gap correlated positively with years of occupational exposure (*r* = 0.67; *p* < 0.001). Logistic regression identified wrist ulnar deviation angle (OR = 2.14; 95% CI 1.43-3.19) and grip force asymmetry (OR = 1.87; 95% CI 1.21-2.89) as independent predictors of instability signs.

**Conclusions:** Manual laborers with work-related scapholunate instability signs exhibit distinct biomechanical signatures during dynamic gripping, including reduced force output, abnormal wrist posture, and compensatory neuromuscular recruitment. Occupational exposure duration is a significant predictor of radiographic scapholunate dissociation. These findings support the integration of ergonomic risk screening and early biomechanical assessment into occupational health protocols for manual labor populations in low- and middle-income settings.

## INTRODUCTION

Work-related musculoskeletal disorders (WMSDs) of the upper extremity constitute a major global occupational health burden, disproportionately affecting manual laborers engaged in construction, manufacturing, and heavy industry (1, 2). Among wrist pathologies, scapholunate instability (SLI) is the most frequently encountered form of carpal instability and is characterized by disruption or attenuation of the scapholunate interosseous ligament (SLIL), resulting in abnormal kinematic coupling between the scaphoid and lunate bones (3, 4). While acute SLI is classically attributed to a fall on an outstretched hand, emerging evidence suggests that chronic, cumulative occupational loading — through repetitive forceful gripping, sustained axial compression, and awkward wrist postures — may represent an underappreciated pathomechanical pathway to progressive ligamentous compromise (5, 6).

The scapholunate joint occupies a pivotal functional role in force transmission across the carpus. Under physiological loading, the SLIL maintains synchronous rotation between the scaphoid and lunate, distributing compressive forces from the radius through the proximal carpal row to the distal row during grip activities (7, 8). Disruption of this linkage produces pathological scaphoid flexion and lunate extension — the DISI (dorsal intercalated segment instability) deformity — which substantially reduces the wrist’s capacity to sustain functional loads (4, 9). In manual laborers, gripping forces routinely exceed 300 N during occupational tasks such as hammer swinging, pipe wrenching, and shovel handling (10), and are often generated in positions of combined wrist extension and ulnar deviation that may impose pathological torque on the SLIL (11).

Despite the prevalence of grip-intensive occupations and the known biomechanical vulnerability of the scapholunate complex under dynamic loading, relatively few studies have systematically characterized the grip biomechanics and wrist kinematics of symptomatic manual laborers. Existing literature is predominantly derived from cadaveric or imaging-based models (12, 13).

Understanding the specific dynamic grip force profiles, wrist motion patterns, and neuromuscular compensatory strategies present in manual laborers with SLI signs is essential for both clinical diagnosis and ergonomic intervention. Such data can inform targeted workplace modifications, early screening protocols, and rehabilitation programs that may prevent progression from partial SLIL injury to scapholunate advanced collapse (SLAC) arthritis — the end-stage consequence of untreated instability (4, 14).

The purpose of the present study was to quantify and compare dynamic grip force profiles and wrist kinematics between manual laborers exhibiting clinical SLI signs and asymptomatic worker controls, to characterize associated neuromuscular recruitment patterns using surface electromyography, to identify occupation-related biomechanical risk predictors of SLI signs in this population. We hypothesized that symptomatic workers would demonstrate significantly reduced grip force, greater wrist ulnar deviation during peak grip, and compensatory hyperactivation of wrist flexors compared with controls.

## METHODS

### Study Design and Ethical Approval

A cross-sectional observational study was conducted between December 2025 and June 2026 in Sialkot District, Punjab, Pakistan. The study protocol was reviewed and approved by the institutional review board of Islam College of Physical Therapy, Sialkot. All participants provided written informed consent prior to enrollment. The study adhered to the ethical principles of the Declaration of Helsinki.

### Participant Recruitment and Eligibility

Adult male manual laborers aged 20–55 years with a minimum of three years of continuous employment in construction, civil engineering, or heavy manufacturing were eligible for inclusion. Participants were recruited from six work sites in Sialkot District via purposive sampling following workplace coordination with site supervisors.

### Inclusion Criteria

1. Positive Watson scaphoid shift test with reproduced dorsal wrist pain on the dominant side.
2. Localized scapholunate interval tenderness on dorsal palpation.
3. Self-reported wrist pain during gripping for ≥ 8 weeks.
4. Scapholunate gap ≥ 3 mm on posteroanterior radiograph with clenched-fist view. Participants in the control group were matched workers from the same occupational settings who were asymptomatic, had no history of wrist injury or surgery, and had a negative Watson test bilaterally.

### Exclusion Criteria

1. Prior wrist fracture or surgical intervention
2. Inflammatory arthritis or connective tissue disease
3. Peripheral neuropathy; pregnancy
4. Inability to perform standardized gripping tasks due to any cause
5. Radiographic evidence of significant wrist osteoarthritis (Kellgren–Lawrence grade ≥ 2).

### Sample Size

We performed a prospective calculation of the needed sample size with the G Power software(Version 3.1.9.7) for an independent samples t-test, assuming a large effect size (Cohen’s d = 0.90) based on grip force differences reported in comparable occupational wrist studies, 80% statistical power, and α = 0.05. The minimum required sample was 18 per group; to account for potential attrition of 15%, 21 participants per group were enrolled, yielding a total of 42 participants.

### Clinical Assessment

#### Watson Scaphoid Shift Test

Stabilizing the scaphoid tubercle on the palmar surface while passively moving the wrist from ulnar to radial deviation. A positive result was defined as a palpable or audible clunk with reproduction of dorsal wrist pain, confirmed on both initial and repeat assessment with comparison to the contralateral wrist (15, 16).

#### Radiographic Evaluation

Posteroanterior wrist radiographs were obtained in the neutral and clenched-fist positions. Scapholunate gap, scapholunate angle, and radiolunate angle were measured by an independent musculoskeletal radiologist blinded to group allocation. A scapholunate angle >70degrees was considered indicative of DISI deformity (4).

### Dynamic Grip Force Assessment

Grip force was measured using a calibrated Jamar hydraulic hand dynamometer (Lafayette Instrument Company, Lafayette, IN, USA). Participants were seated with the shoulder adducted, elbow flexed to 90degrees, forearm in the neutral position, and wrist in a neutral 0degrees position, consistent with American Society of Hand Therapists (ASHT) standardized protocol. Five maximal voluntary isometric contraction (MVIC) trials were performed, each lasting five seconds, with 60-second rest intervals between trials. The mean of the three middle trials was used for analysis. Grip force asymmetry index was calculated as the absolute force difference between dominant and non-dominant hands divided by the dominant hand force, expressed as a percentage.

### Wrist Kinematics

Wrist kinematics during a standardized simulated gripping task were recorded using a biaxial electrogoniometer, model no. SG65 (Biometrics Ltd., Newport, UK) affixed to the dorsum of the hand and forearm. The simulated gripping task required participants to grasp and lift a standardized 5 kg cylindrical load (diameter 40 mm) from a bench, mimicking a common occupational handling pattern. Joint angles in the flexion-extension and radial-ulnar deviation planes were sampled at 1000 Hz. Peak wrist ulnar deviation angle, angular velocity, and range of motion during the dynamic gripping cycle were extracted for analysis.

### Surface Electromyography

Surface EMG was recorded from three forearm muscles: flexor digitorum superficialis (FDS), flexor carpi ulnaris (FCU), and extensor carpi radialis longus (ECRL), using disposable Ag/AgCl electrodes (Ambu BlueSensor; 20 mm inter-electrode distance) placed following SENIAM guidelines. Signals were amplified (gain 1000×), bandpass filtered (10–450 Hz), and sampled at 2000 Hz using a 16-channel EMG system (Noraxon Ultium; Noraxon USA Inc., Scottsdale, AZ, USA). Raw EMG was rectified and smoothed with a 100 ms root mean square window. Amplitude was normalized to the corresponding MVIC value to yield percentage of maximum voluntary contraction (%MVC) for between-subject comparisons.

### Statistical Analysis

All data was assessed for normality using the Shapiro-Wilk test. Descriptive statistics are presented as mean standard deviation (SD) or median [interquartile range] as appropriate. Independent samples t-tests were used to compare normally distributed continuous variables between groups. Where normality was violated, the Mann-Whitney U test was applied. Pearson correlation was used to assess relationships between radiographic scapholunate gap and occupational exposure (years). Binary logistic regression was performed to identify independent biomechanical predictors of SLI clinical signs, with results expressed as odds ratios (OR) and 95% confidence intervals (CI). The significance threshold was set at α = 0.05. Statistical analyses were performed in IBM SPSS Statistics version 28.0.

## RESULTS

### Participant Characteristics

Forty-two participants were enrolled, with 21 in the SLI group and 21 in the control group. No participants were lost to follow-up or excluded post-enrollment. Demographic and occupational characteristics are summarized in

Table 1. The two groups were well-matched for age (SLI: 36.4 SD 7.2 years; control: 34.8 SD 6.9 years; p = 0.46), BMI (SLI: 24.7 SD 2.9 kg/m^2^; control: 24.3 SD 3.1 kg/m^2^; p = 0.64), and dominant hand (all right-handed). Mean occupational exposure was significantly greater in the SLI group (12.3 SD 3.8 years) compared with controls (7.4 SD 2.9 years; *p* < 0.001).

**Table 1.** Demographic and Occupational Characteristics of Study Participants.

| Characteristic | SLI Group (n = 21) | Control Group (n = 21) | p Value |
| --- | --- | --- | --- |
| Age (years) | 36.4 SD 7.2 | 34.8 SD 6.9 | 0.46 |
| BMI (kg/m<sup>2</sup>) | 24.7 SD 2.9 | 24.3 SD 3.1 | 0.64 |
| Occupational exposure (years) | 12.3 SD 3.8 | 7.4 SD 2.9 | < 0.001 |
| Primary occupation:<br>Construction (%) | 71.4 | 66.7 | 0.73 |
| Primary occupation:<br>Manufacturing (%) | 28.6 | 33.3 | 0.73 |
| Dominant hand: Right<br>(%) | 100 | 100 | — |
| Scapholunate gap, mm | 3.7 SD 0.6 | 1.6 SD 0.4 | < 0.001 |
| Scapholunate angle<br>(degrees) | 74.2 SD 8.3 | 52.4 SD 6.7 | < 0.001 |

Values are mean SD unless otherwise stated. BMI, body mass index; SLI, scapholunate instability. p values from independent samples t-test (continuous) or chi-square test (categorical).

### Dynamic Grip Force

Dominant hand grip force was significantly lower in the SLI group than in controls (28.4 SD 5.6 kg versus 41.7 SD 6.3 kg; mean difference 13.3 kg; 95% CI 9.8–16.8; *p* < 0.001; Cohen’s d = 2.25). The grip force asymmetry index was also significantly greater in the SLI group (18.9 SD 6.2% versus 7.3 SD 3.1%; *p* < 0.001). These data are presented in Table 2.

**Table 2.** Dynamic Grip Force and Asymmetry Index by Group.

| Variable | SLI Group | Control Group | Mean Difference (95% CI) | p Value |
| --- | --- | --- | --- | --- |
| Dominant grip force (kg) | 28.4 SD 5.6 | 41.7 SD 6.3 | 13.3 (9.8–16.8) | < 0.001 |
| Non-dominant grip force (kg) | 35.1 SD 5.9 | 40.2 SD 5.8 | 5.1 (2.0–8.2) | 0.002 |
| Grip force asymmetry index (%) | 18.9 SD 6.2 | 7.3 SD 3.1 | 11.6 (8.4–14.8) | < 0.001 |
| Peak grip rate of force development (N/s) | 142.6 SD 38.4 | 198.3 SD 41.7 | 55.7 (33.1–78.3) | < 0.001 |

Values are mean SD SD. CI, confidence interval; SLI, scapholunate instability.

### Wrist Kinematics During Dynamic Gripping

During the simulated occupational gripping task, the SLI group exhibited significantly greater peak wrist ulnar deviation (22.3degrees SD 4.1degrees versus 14.6degrees SD 3.8degrees; *p* = 0.002) and a greater wrist angular velocity during the loading phase (48.7 SD 9.2degrees/s versus 36.4 SD 8.6degrees/s; *p* = 0.003). Wrist flexion-extension range of motion during the grip cycle did not differ significantly between groups (SLI: 41.2degrees SD 8.6degrees; control: 39.7degrees SD 7.4degrees; *p* = 0.54). Kinematic data are presented in Table 3.

**Table 3.** Wrist Kinematic Parameters During Standardized Dynamic Gripping Task.

| Kinematic Variable | SLI Group | Control Group | *p* Value |
| --- | --- | --- | --- |
| Peak ulnar deviation (degrees) | 22.3 SD 4.1 | 14.6 SD 3.8 | 0.002 |
| Peak flexion during load (degrees) | 18.6 SD 5.2 | 17.9 SD 4.8 | 0.63 |
| Angular velocity, loading phase (degrees/s) | 48.7 SD 9.2 | 36.4 SD 8.6 | 0.003 |
| Flexion-extension ROM (degrees) | 41.2 SD 8.6 | 39.7 SD 7.4 | 0.54 |
| Radial-ulnar deviation ROM (degrees) | 28.4 SD 5.7 | 22.1 SD 4.9 | 0.01 |

Values are mean SD SD. ROM, range of motion; SLI, scapholunate instability.

### Surface Electromyography

Compared with controls, the SLI group demonstrated significantly higher FDS activation during the peak grip phase (62.3 SD 10.4% MVC versus 44.1 SD 8.7% MVC; *p* < 0.001) and FCU activation (44.7 SD 9.8% MVC versus 31.2 SD 7.6% MVC; p = 0.001). ECRL co-activation was also elevated in the SLI group (38.6 SD 8.2% MVC versus 28.9 SD 6.4% MVC; *p* = 0.004), suggesting a pattern of global wrist muscle hyperactivation as a neuromuscular stabilization strategy. EMG findings are presented in Table 4.

**Table 4.** Surface EMG Amplitude (%MVC) During Dynamic Gripping by Group.

| Muscle | SLI Group<br>(%MVC) | Control Group<br>(%MVC) | p<br>Value |
| --- | --- | --- | --- |
| Flexor digitorum superficialis<br>(FDS) | 62.3 SD 10.4 | 44.1 SD 8.7 | <<br>0.001 |
| Flexor carpi ulnaris (FCU) | 44.7 SD 9.8 | 31.2 SD 7.6 | 0.001 |
| Extensor carpi radialis longus<br>(ECRL) | 38.6 SD 8.2 | 28.9 SD 6.4 | 0.004 |

Values are mean SD. %MVC, percentage maximum voluntary contraction; ECRL, extensor carpi radialis longus; FCU, flexor carpi ulnaris; FDS, flexor digitorum superficialis; SLI, scapholunate instability.

### Correlation and Regression Analyses

Scapholunate gap on radiograph correlated positively and significantly with years of occupational exposure in the SLI group (Pearson *r* = 0.67; *p* < 0.001), suggesting cumulative biomechanical loading as a driver of carpal dissociation. Binary logistic regression identified two independent predictors of clinical SLI signs: peak wrist ulnar deviation during gripping (OR = 2.14; 95% CI 1.43–3.19; *p* < 0.001) and dominant hand grip force asymmetry index (OR = 1.87; 95% CI 1.21–2.89; *p* = 0.005), after controlling for age and occupational exposure duration. The overall model was statistically significant (*Chi-square* = 34.7; df = 4; *p* < 0.001) with a Nagelkerke *R^2^* of 0.72.

## DISCUSSION

This study simultaneously characterize dynamic grip force profiles, wrist kinematics, and neuromuscular recruitment patterns in a cohort of manual laborers exhibiting clinical signs of work-related scapholunate instability. The findings collectively support a coherent biomechanical model in which occupational dynamic gripping under pathological wrist postures generates abnormal torques on the SLIL, leading to progressive kinematic dissociation, reduced force-generating capacity, and compensatory neuromuscular hyperactivation.

Reduction in dominant hand grip force observed in the SLI group (28.4 versus 41.7 kg) is consistent with previous work demonstrating that carpal instability substantially impairs the wrist’s ability to sustain physiological loads due to disrupted kinematic coupling (4, 7).

Watson and colleagues established that scapholunate dissociation fundamentally compromises load transmission through the carpus by allowing the scaphoid to flex independently of the lunate, dissipating force that would otherwise be transmitted through an intact proximal carpal row (4). Our data extend this observation to the occupational context, demonstrating that even at the pre-surgical, clinically detected stage, grip force deficits are large (Cohen’s d = 2.25) and functionally meaningful — exceeding the 10 kg minimal detectable change threshold for occupational performance impairment described in hand surgery rehabilitation literature (17).

The finding that peaked wrist ulnar deviation during gripping was significantly greater in the SLI group (22.3degrees versus 14.6degrees) is mechanistically important. Cadaveric studies have demonstrated that SLIL torque increases markedly as the wrist adopts ulnar deviation combined with loading (11, 18). In the occupational setting, tasks such as hammer swinging, wrench tightening, and repetitive lifting in ulnar-deviated wrist postures are ubiquitous, particularly among construction workers — a population comprising 71% of our SLI group.

This postural loading pattern is distinct from the classic traumatic mechanism (fall on outstretched hand with forced extension and radial deviation) and may represent a chronic, low-grade injury mechanism that progressively attenuates the SLIL dorsal fibers, which provide the primary torsional restraint (8, 12).

The sEMG data revealed paradoxical hyperactivation of wrist flexors (FDS, FCU) and wrist extensors (ECRL) in the SLI group. This pattern is consistent with a compensatory neuromuscular co-activation strategy in which muscles are recruited to dynamically stabilize an unstable joint, substituting for passive ligamentous restraint (19).

Similar compensatory EMG patterns have been described in patients with anterior cruciate ligament deficiency and glenohumeral instability, where periarticular muscle co-activation increases in proportion to the degree of passive restraint loss (20). In the wrist, the wrist flexor-extensor muscle complex has been proposed to serve as an important dynamic stabilizer of the scapholunate interval (21), and our findings suggest that manual laborers with SLI signs actively exploit this mechanism. However, sustained muscle co-activation imposes elevated metabolic demands on the forearm musculature, which may predispose workers to secondary fatigue-related injury and accelerate SLIL degeneration over the occupational career.

The strong positive correlation between radiographic scapholunate gap and years of occupational exposure (r = 0.67; p < 0.001) provides novel epidemiological evidence supporting a dose-response relationship between cumulative manual labor exposure and carpal dissociation. This is congruent with a growing body of occupational health literature demonstrating that work-related wrist disorders follow an exposure-dependent trajectory, with forceful repetitive gripping, sustained wrist loading, and hand-arm vibration identified as primary risk factors (1, 6, 22). Our logistic regression identified peak wrist ulnar deviation and grip force asymmetry as independent predictors of SLI clinical signs, underscoring the diagnostic utility of these biomechanical measures in occupational screening contexts.

Strengths of this study include the use of objectively validated biomechanical instrumentation, comprehensive multi-domain assessment (force, kinematics, EMG, and imaging), a well-matched control group drawn from the same occupational cohort, and conduct within an under-researched low-middle-income country (LMIC) occupational setting. Limitations include the cross-sectional design, which precludes causal inference; the single-sex male cohort, limiting generalizability to female manual laborers; the absence of MRI for definitive SLIL grading, relying instead on clinical and radiographic criteria; and the use of simulated rather than actual in situ occupational gripping tasks. Future longitudinal cohort studies incorporating MRI-based SLIL grading and in-field biomechanical monitoring are warranted to establish causality and optimal surveillance intervals.

Clinically, these findings have direct implications for occupational physiotherapy practice in Pakistan. The identification of wrist ulnar deviation angle and grip asymmetry as predictors of SLI signs suggests that simple, low-cost screening measures including goniometric wrist posture observation during work tasks and bilateral Jamar dynamometry could be integrated into routine occupational health assessments in manual-labor environments. Early detection of SLI signs may enable targeted ergonomic modifications (tool redesign, wrist splinting during high-risk tasks, job rotation) and physiotherapy intervention before progression to irreversible SLAC arthritis.

## CONCLUSION

Manual laborers exhibiting clinical signs of work-related scapholunate instability demonstrate a distinctive biomechanical signature during dynamic gripping: reduced grip force output, elevated wrist ulnar deviation, and compensatory neuromuscular co-activation of wrist flexors and extensors. Scapholunate gap is positively associated with cumulative occupational exposure, and peak wrist ulnar deviation during gripping emerges as the strongest independent biomechanical predictor of clinical instability signs. These findings support the development of biomechanically informed occupational screening protocols and ergonomic interventions in manual labor populations, particularly in LMIC settings where access to advanced diagnostic imaging may be limited.

## Data Availability

All data produced in the present study are available upon reasonable request to the authors

## Conflict of Interest

The authors have declared no conflict of interest.

## Funding

This research received no specific grant from any funding agency in the public, commercial, or not-for-profit sectors.

## Abbreviations

CI: confidence interval
DRUJ: distal radioulnar joint
EMG/sEMG: (surface) electromyography
FCU: flexor carpi ulnaris
FDS: flexor digitorum superficialis
MSD: musculoskeletal disorder
OR: odds ratio
ROM: range of motion
SL/SLI: scapholunate/scapholunate instability
SLIL: scapholunate interosseous ligament
SLAC: scapholunate advanced collapse
WMSDs: work-related musculoskeletal disorders
SD: standard deviation

## Notes

### Competing Interest Statement

The authors have declared no competing interest.

### Author Declarations

Ethics committee/IRB of Islam College of Physical Therapy gave ethical approval for this work.

## References

1. Rodríguez-Pulido AG, Arrieta-Córdova AF, Arce-Huamani MA. Prevalence and correlation of workload and musculoskeletal disorders in industrial workers: a cross-sectional study. Frontiers in Rehabilitation Sciences. 2025;Volume 6 - 2025.

2. Tolera ST, Gobena T, Geremew A, Toseva E, Assefa N. Work-related musculoskeletal disorders and associated factors among hospital sanitary workers in public hospitals of Eastern Ethiopia. BMC Musculoskelet Disord. 2025;26(1):640.

3. Wessel L, Wolfe S. Scapholunate Instability: Current Concepts in Diagnosis and Management Classification and Treatment Considerations-Part 2. The Journal of hand surgery. 2023;48.

4. Watson HK, Ballet FL. The SLAC wrist: scapholunate advanced collapse pattern of degenerative arthritis. J Hand Surg Am. 1984;9(3):358–65.

5. Palmer AK, Werner FW. Biomechanics of the distal radioulnar joint. Clin Orthop Relat Res. 1984(187):26–35.

6. Sluiter JK, Rest KM, Frings-Dresen MH. Criteria document for evaluating the work-relatedness of upper-extremity musculoskeletal disorders. Scand J Work Environ Health. 2001;27 Suppl 1:1–102.

7. Short WH, Werner FW, Green JK, Masaoka S. Biomechanical evaluation of ligamentous stabilizers of the scaphoid and lunate. J Hand Surg Am. 2002;27(6):991–1002.

8. Berger RA. The gross and histologic anatomy of the scapholunate interosseous ligament. J Hand Surg Am. 1996;21(2):170–8.

9. Linscheid RL, Dobyns JH, Beabout JW, Bryan RS. Traumatic instability of the wrist. Diagnosis, classification, and pathomechanics. J Bone Joint Surg Am. 1972;54(8):1612–32.

10. Bhattacharya A, McGlothlin JD. Occupational ergonomics. Theory and applications: Marcel Dekker; 1996.

11. Garcia-Elias M, An KN, Cooney WP, 3rd, Linscheid RL, Chao EY. Stability of the transverse carpal arch: an experimental study. J Hand Surg Am. 1989;14(2 Pt 1):277–82.

12. Mitsuyasu H, Patterson RM, Shah MA, Buford WL, Iwamoto Y, Viegas SF. The role of the dorsal intercarpal ligament in dynamic and static scapholunate instability. J Hand Surg Am. 2004;29(2):279–88.

13. Kaufmann RA, Pfaeffle HJ, Blankenhorn BD, Stabile K, Robertson D, Goitz R. Kinematics of the midcarpal and radiocarpal joint in flexion and extension: an in vitro study. J Hand Surg Am. 2006;31(7):1142–8.

14. Ruch DS, Poehling GG. Arthroscopic management of partial scapholunate and lunotriquetral injuries of the wrist. J Hand Surg Am. 1996;21(3):412–7.

15. Watson HK, Ashmead Dt, Makhlouf MV. Examination of the scaphoid. J Hand Surg Am. 1988;13(5):657–60.

16. Schmauss D, Pöhlmann S, Weinzierl A, Schmauss V, Moog P, Germann G, et al. Relevance of the Scaphoid Shift Test for the Investigation of Scapholunate Ligament Injuries. J Clin Med. 2022;11(21).

17. Mathiowetz V, Kashman N, Volland G, Weber K, Dowe M, Rogers S. Grip and pinch strength: normative data for adults. Arch Phys Med Rehabil. 1985;66(2):69–74.

18. Patterson RM, Nicodemus CL, Viegas SF, Elder KW, Rosenblatt J. High-speed, three-dimensional kinematic analysis of the normal wrist. J Hand Surg Am. 1998;23(3):446–53.

19. Mannella K, Forman GN, Mugnosso M, Zenzeri J, Holmes MWR. The effects of isometric hand grip force on wrist kinematics and forearm muscle activity during radial and ulnar wrist joint perturbations. PeerJ. 2022;10:e13495.

20. Solomonow M, Baratta R, Zhou BH, Shoji H, Bose W, Beck C, et al. The synergistic action of the anterior cruciate ligament and thigh muscles in maintaining joint stability. Am J Sports Med. 1987;15(3):207–13.

21. Wolfe SW, Crisco JJ, Orr CM, Marzke MW. The dart-throwing motion of the wrist: is it unique to humans? J Hand Surg Am. 2006;31(9):1429–37.

22. Larsen CF, Amadio PC, Gilula LA, Hodge JC. Analysis of carpal instability: I. Description of the scheme. The Journal of Hand Surgery. 1995;20(5):757–64.

